# Double burden of malnutrition and its social disparities among rural Sri Lankan adolescents

**DOI:** 10.1101/2022.03.29.22273121

**Authors:** B. L. Goonapienuwala, N. S. Kalupahana, S. B. Agampodi, S. Siribaddana

**Affiliations:** Department of Physiology, Faculty of Medicine and Allied Sciences, Rajarata University of Sri Lanka, Saliyapura, Sri Lanka; Department of Physiology, Faculty of Medicine, University of Peradeniya, Peradeniya, Sri Lanka; Department of Community Medicine, Faculty of Medicine and Allied Sciences, Rajarata University of Sri Lanka, Saliyapura, Sri Lanka; Department of Medicine, Faculty of Medicine and Allied Sciences, Rajarata University of Sri Lanka, Saliyapura, Sri Lanka

## Abstract

In Sri Lanka, the double burden of nutrition is often neglected, and increase of adolescent obesity is not well investigated. This study determines the double burden of malnutrition among adolescents in Anuradhapura district exploring the differences of prevalence based on different definitions. Students aged 13 to 16 years were selected from 74 schools using probability proportionate to size sampling. Anthropometry was done according to WHO guidelines. Obesity was defined according to body mass index (BMI) based definitions of WHO, International Obesity Task Force and Indian growth references. Central obesity was defined using Indian and British waist circumference cut-offs. Prevalence estimates from different definitions were compared using McNemar’s test. Socio-demographic determinants of nutritional issues were assessed using Chi-square test for independence. A total of 3105 students (47.7% boys) were studied (mean age 14.8± 0.8 years). According to WHO definitions, 73 (2.4%, 95% CI; 1.9–2.9) were obese, 222 (7.2%, 95% CI; 6.3–8.1) were overweight, 673 (21.7%, 95% CI; 20.2–23.1,) were thin and 396 (12.8%, 95% CI; 11.6–14.0) were stunted. More boys (3.1%) than girls (1.7%) were obese as well as thin (29.0% compared to 15.0%). Prevalence of overweight/obesity was higher among students in larger, urban schools, and belonging to high social class and more educated parents. Prevalence of overweight/obesity estimated using IOTF-Asian and Indian thresholds were significantly higher than that from WHO and IOTF-international thresholds. Double burden of malnutrition is affecting the adolescents in rural Sri Lanka. Prevalence estimates of obesity largely depend on the definition used.

## Introduction

The mean age adjusted body mass index (BMI) for children and adolescents has risen from 17.2 to 18.6 among girls and 16.8 to 18.5 among boys from 1975 to 2016 [1]. As a result, the prevalence of overweight and obesity has increased from 8.1% to 12.9% among boys and 8.4% to 13.4% among girls in low and middle-income countries. The respective prevalence in high-income countries was 23.8% and 22.6% [2]. This increase has a direct contribution to more than 17 million deaths due to cardiovascular diseases [3] as well as global morbidity and mortality due to many other non-communicable diseases worldwide.

Adolescents make up about 18% of the world population and more than half of those live in Asia [4]. Since adolescents are typically considered a “healthy” group, less attention has been paid on adolescent health. The adolescent health programs are traditionally focusing on reproductive and sexual health issues. In regions such as South and Central Asia, where the under-nutrition rates are still high, the double burden of nutrition is often neglected, and the increase of childhood and adolescent obesity is not well addressed. Even in places where studies are carried out, the focus is on urban adolescents. Further, the measurement diversity as proposed by different international agencies (WHO, NHANES III, IOTF, CDC) makes the comparison difficult.

Obesity and overweight can be assessed using several measures; e.g., body mass index (BMI), waist circumference (WC), waist to hip ratio, waist to height ratio (WHtR) and skin fold thickness. BMI is considered the best and most widely used anthropometric index in population studies but sensitivity to detect obesity is questioned [5,6]. Although there is a good correlation between BMI and body fat, there is evidence for ethnic differences in body fat content. Asians have higher body fat percentage compared to Caucasians for a given BMI [7]. These led to development of country specific thresholds in many countries including USA, UK and India.

In Sri Lanka, research on adolescent nutrition was mainly conducted in urban areas. Prevalence of overweight and obesity in Colombo was between 1.9% and 13% among schooling adolescents [6,8–11]. The wide variation in estimates is due to different definitions used as well as selection biases in some studies. Due to the higher genetic risk for cardiovascular diseases, local thresholds for BMI and WC [12] for Sri Lankan children have been proposed recently. However, due to lack of consensus, international definitions are still being used in most studies. The present study aimed to determine the prevalence of obesity and the existence of double burden of malnutrition in rural Sri Lanka, eliminating the sampling biases to provide better estimates as well as to explore the discrepancies with different definitions to detect obesity.

## Materials and Methods

### Design, setting and population

This school based cross sectional descriptive study was carried out in Sinhala medium schools in Anuradhapura district, Sri Lanka, from April 2013 to November 2014. In the district, 94% of the population was rural and 90.9% were Sinhalese [13]. There were 546 schools in the district and 481 of them were Sinhala medium. Of this, 322 schools had classes up to grade11 or above [14]. There were total of 45,391 students with 12,175 in grade 9, 12,150 in grade 10 and 21,066 in grade 11.

### Study sample and sampling technique

The sample size was calculated to detect 5% prevalence of overweight and obesity with 0.2 precision with 95% confidence interval. Sample size was adjusted for design effect of 1.5 because of cluster sampling. Total calculated sample size was 3036. The cluster size was the most frequent number of students (33 students) per class and there were 92 clusters (3036/33). Cluster selection was done using probability proportionate to size sampling and the 92 clusters were finally drawn from 74 schools.

### Study instruments and measurements

Anthropometric measurements were carried out according to WHO guidelines [15]. Height was measured to the nearest millimeter using portable stadiometers (Seca 213 – Germany); standing without footwear, with heel, back and occiput touching the measuring board and eyes at the same level as ears (Frankfurt plane). Weight was measured to the nearest 100g with student wearing the uniform and without footwear, using portable digital weighing scales (Seca 803 – Germany). WC was measured to the nearest millimeter using a flexible tape at the midpoint between the lowest rib and the superior iliac spine during expiration [15]. Six pre-registration medical graduates were trained on anthropometric measurements and data collection. Observer and instrument error for the measurements were assessed on nine students with the six observers trained on anthropometric measurements using two instruments for each measurement. Separate height and weight measuring scales were used for girls and boys and the same scales were used throughout the study. The two waist measuring tapes were regularly compared with a standard tape for discrepancies in calibration. When the discrepancy was found to be more than 1 mm, that tape was replaced with a new one.

Waist circumference was measured over the school uniform and without the uniform as a preliminary study and correction factors were applied for WC measurements taken over the uniform. Each school was visited twice. During the first visit, classes were selected and consent forms for parents and information leaflets were distributed. Second visit was to collect data. Students completed the self-administered questionnaire on socio-demographic data. Each anthropometric measurement was repeated thrice, and the mean value was taken.

### Analysis of data

Age and sex segregated descriptive statistics were carried out for height, weight, WC, BMI, WHtR, z-scores of BMI for age and height for age. Z-Scores for BMI and height were calculated using WHO Anthro-plus software [16]. Thinness (BMI for age less than 2SD), severe thinness (BMI for age less than 3SD), stunting (height for age less than 2SD) and severe stunting (height for age less than 3SD) were defined according to WHO 2007 growth references [17]. Hereafter, unless specified, “thinness” also includes those with severe thinness, and “stunting” includes those with severe stunting.

We used following definitions of obesity in our study.

1. WHO child growth references [17]

Based on BMI z-score for age; overweight > 1SD but equals or less than 2SD and obese > 2SD. This is for children and adolescents aged 5 to 19 years. Overweight category does not include individuals with obesity.

2. International Obesity Task Force (IOTF) [18]

BMI thresholds for overweight and obesity for each month from two to 18 years of age are available in the IOTF database. We used both Asian and international BMI thresholds.

3. Proposed Indian BMI thresholds [19]

Indian Academy of Pediatrics has given age and sex specific BMI thresholds for overweight and obesity for every six months from five to 18 years.

Visceral obesity was again estimated using different indices;

1. 70^th^ percentile of WC for Indian children [20]

A study involving five major cities covering India has defined the 70^th^ percentile of WC as the threshold for diagnosing metabolic syndrome in Indian children.

However, this study measured the WC in accordance with the NHANES methodology (above upper lateral border of the right ilium). A prediction equation was used to convert the WC according to WHO anthropometric guidelines [21].

2. 90^th^ percentile of WC for British children [22]

International Diabetes Federation recommended the 90^th^ percentile of WC to define central obesity as one of the diagnostic criteria of metabolic syndrome in children [23].

3. 75^th^ percentile of WC for south Indian children [24]

A South Indian study has suggested the use of 75^th^ percentile from their study as an action point in identifying obesity in Indian children.

For the purpose of analysis, parental education level was divided into two categories (educated up to grade 11, and above GCE (O/L) exam level). Social class was determined on the highest-ranking occupation of the parents.

Data were analyzed using IBM SPSS 20^th^ version.

### Ethical concerns

Ethical approval of the study was obtained from the Ethics review committee of the faculty of Medicine and Allied Sciences, Rajarata University of Sri Lanka. Permission was obtained from the provincial and zonal education departments and principals of the selected schools. Informed written consent was taken from the parents and students for participation in the study as well as possible publication of the results. Anonymity was maintained throughout data handling process. A health education program was organized for those who were detected as overweight and obese. They were referred for further medical management depending on the results of screening tests for metabolic derangements.

## Results

A total of 3135 students were recruited and age specific growth parameters were available for 3105 (99.0%) students. Of this 3105, 1481 (47.7%) were boys and 3094 (99.6%) were Sinhalese. Mean age (SD) of the sample was 14.8 (0.8) years (range = 13.0 to 16.9). Table 1 shows the characteristics of the study sample.

**Table 1:** Socio-demographic characteristics of the 3105 adolescents from rural Sri Lanka included in the study sample.

|  | **Boys** | **Girls** |
| --- | --- | --- |
| Number of students | 1481 (47.7) | 1624 (52.3) |
| Mean age (years) | 14.8 (SD = 0.8) | 14.8 (SD =0.8) |
| Ethnicity |  |  |
| Sinhala | 1471 (99.3) | 1623 (99.9) |
| Tamil | 0 (0.0) | 0 (0.0) |
| Moor/Malay | 9 (0.6) | 1 (0.1) |
| Other | 1 (0.1) | 0 (0.0) |
| Grade in school |  |  |
| 9 | 496 (33.7) | 515 (31.8) |
| 10 | 712 (48.4) | 830 (51.2) |
| 11 | 262 (17.8) | 276 (17.0) |
| Education zone |  |  |
| Anuradhapura | 541 (36.5) | 532 (32.8) |
| Kekirawa | 236 (15.9) | 270 (16.6) |
| Tambuththegama | 307 (20.7) | 353 (21.7) |
| Galenbindunuwewa | 182 (12.3) | 225 (13.9) |
| Kebithigollewa | 215 (14.5) | 244 (15.0) |
| School type |  |  |
| 1AB | 568 (38.4) | 586 (36.1) |
| 1C | 580 (39.2) | 635 (39.1) |
| Type 2 | 333 (22.5) | 403 (24.8) |
| Mother's education level |  |  |
| Up to grade 11 | 740 (66.8) | 862 (65.3) |
| O/L and above | 368 (33.2) | 458 (34.7) |
| Father's education level |  |  |
| Up to grade 11 | 697 (67.0) | 762 (64.7) |
| O/L and above | 344 (33.0) | 415 (35.3) |
| Employed mothers | 518 (35.3) | 567 (35.3) |
| Employed fathers | 1351 (92.9) | 1463 (92.7) |
| Social class |  |  |
| High | 100 (7.3) | 98 (6.4) |
| Middle | 1160 (84.1) | 1265 (82.1) |
| Low | 119 (8.6) | 178 (11.6) |

Number of boys and girls in each category is given along with percentage within parenthesis.

### Distribution of anthropometric measurements and nutritional status

Table 2 shows age and sex segregated distribution of anthropometry, BMI–for-age and height-for-age Z scores. Distribution of severe thinness, thinness, overweight, obesity, severe stunting and stunting in the study sample are also shown.

**Table 2:** Age and sex segregated distribution of growth parameters among schooling adolescents in the Anuradhapura district, Sri Lanka.

|  | 13 Years<sup>a</sup> |  | 14 Years |  | 15 Years |  | 16 Years |  |
| --- | --- | --- | --- | --- | --- | --- | --- | --- |
|  | Boys<br>(n= 186) | Girls<br>(n =225) | Boys<br>(n=574) | Girls<br>(n=639) | Boys<br>(n=578) | Girls<br>(n=608) | Boys<br>(n=143) | Girls<br>(n=152) |
| **Mean (SD) of growth parameters** |  |  |  |  |  |  |  |  |
| Height (cm) | 153.5 (7.6)<sup>b</sup> | 151.4 (6.0) | 158.4 (8.1)<sup>b</sup> | 153.3 (5.5) | 162.6 (7.4)<sup>b</sup> | 154.6 (5.4) | 164.4 (5.8)<sup>b</sup> | 153.8 (5.4) |
| Weight (kg) | 41.7 (11.6) | 40.6 (8.1) | 44.4 (11.0) | 43.8 (8.5) | 48.6 (10.8)<sup>b</sup> | 45.1 (8.2) | 50.9(10.4)<sup>b</sup> | 45.7 (7.8) |
| Waist circumference (cm) | 62.3 (9.0)<sup>b</sup> | 60.2 (6.8) | 63.2 (9.0) | 62.3 (7.2) | 65.4 (8.8)<sup>b</sup> | 63.0 (7.0) | 66.4 (8.6)<sup>b</sup> | 63.7 (7.2) |
| BMI (kg/m<sup>2</sup>) | 17.5 (3.7) | 17.7 (3.0) | 17.5 (3.4)<sup>b</sup> | 18.6 (3.2) | 18.3 (3.4)<sup>b</sup> | 18.8 (3.1) | 18.8 (3.1) | 19.3 (3.3) |
| Waist to height ratio | 0.41 (0.1) | 0.40 (0.0) | 0.40 (0.1)<sup>b</sup> | 0.41(0.0) | 0.40 (0.1) | 0.41 (0.0) | 0.40 (0.0) | 0.41 (0.0) |
| Z-Score for BMI | -1.00 (1.6) | -0.93 (1.2) | -1.21 (1.5)<sup>b</sup> | -0.71 (1.2) | -1.10 (1.5)<sup>b</sup> | -0.79 (1.1) | -1.05 (1.3)<sup>b</sup> | -0.73 (1.2) |
| Z-Score for height | -0.92 (1.0) | -1.07 (0.8) | -0.94 (1.0)<sup>b</sup> | -1.07 (0.8) | -0.99 (1.0) | -1.08 (0.8) | -1.17 (0.7) | -1.29 (0.8) |
| **BMI for age & height for age categories - prevalence estimates (with 95% CI)** |  |  |  |  |  |  |  |  |
| Severe thinness<br>< -3 SD | 8.6<br>(5.4–13.5) | 4.4<br>(2.4–8.0) | 11.8<br>(9.5–14.8) | 2.7<br>(1.7–4.2) | 8.1<br>(6.2–10.6) | 1.6<br>(0.9–3.0) | 5.6<br>(2.9–10.6) | 1.3<br>(0.4–4.7) |
| Thinness<br>-3 SD to <-2 SD | 21.5<br>(16.2–28) | 14.7<br>(10.6–19.9) | 19.7<br>(16.6–23.1) | 11.0<br>(8.8–13.6) | 20.2<br>(17.2–23.7) | 12.8<br>(10.4–15.7) | 14.0<br>(9.2–20.6) | 15.8<br>(10.8–22.4) |
| Overweight<br>>+1 SD to +2 SD | 9.1<br>(5.8–14.2) | 4.9<br>(2.8–8.5) | 7.0<br>(5.2–9.4) | 8.5<br>(6.5–10.9) | 8.3<br>(6.3–10.8) | 5.1<br>(3.6–7.1) | 6.3<br>(3.3–11.5) | 7.9<br>(4.6–13.3) |
| Obese<br>> +2 SD | 3.8<br>(1.8–7.6) | 2.2<br>(1.0–5.1) | 3.8<br>(2.5–5.7) | 1.9<br>(1.0–5.1) | 2.4<br>(1.4–4.0) | 1.3<br>(0.7–2.6) | 2.1<br>(0.7–6.0) | 1.3<br>(0.4–4.7) |
| Severe stunting<br><-3SD | 3.8<br>(1.8–7.6) | 2.7<br>(1.2–5.7) | 2.1<br>(1.2–3.6) | 0.6<br>(0.2–1.6) | 2.8<br>(1.7–4.4) | 1.0<br>(0.5–2.1) | 1.4<br>(0.4–5.0) | 0.7<br>(0.1–3.6) |
| Stunting<br>-3 SD to <-2 SD | 8.1<br>(4.9–12.9) | 9.3<br>(6.2–13.8) | 13.6<br>(11.0–16.6) | 10.5<br>(8.4–13.1) | 9.7<br>(7.5–12.4) | 10.9<br>(8.6–13.6) | 10.5<br>(6.5–16.6) | 15.8<br>(10.8–22.4) |

<sup>a</sup> Age group 13 includes 13–13.9 year olds, age group 14 includes 14–14.9 year olds, age group 15 includes 15–15.9 year olds and age group 16
includes 16–16.9 year olds
<sup>b</sup> Difference between boys and girls is statistically significant at 0.05 level

According to WHO growth references, prevalence of age adjusted obesity, overweight, thinness, severe thinness, stunting and severe stunting among boys in this study population were 3.1% (95% CI; 2.3–4.1%, n = 46), 7.7% (95% CI; 6.4–9.2, n = 114), 19.6% (95% CI; 17.6 - 21.7, n = 290), 9.4% (95% CI; 8.0–11.0, n = 139), 11.1% (95% CI; 9.6 - 12.8, n = 164) and 2.5% (95% CI; 1.8–3.4, n = 37) respectively. Among girls, respective values were 1.7% (95% CI; 1.1–2.4%, n = 27), 6.7% (95% CI; 5.5 - 8.0, n = 108), 12.6% (95% CI; 11.1 - 14.3, n = 205), 2.4% (95% CI; 1.8–3.3, n = 39), 11.0% (95% CI; 9.5 - 12.6, n = 178) and 1.0% (95% CI; 0.7–1.7, n = 17). Of the total number of students, 2.4% (95% CI; 1.9–2.9, n = 73) were obese, 7.1% (95% CI; 6.3–8.1, n = 222) were overweight, 15.9% (95% CI; 14.7– 17.3, n = 495) were thin, 5.7% (95% CI; 5.0–6.6, n = 178) were severely thin, 11.0% (95% CI; 9.9–12.2, n = 342) were stunted and 1.7% (95% CI; 1.3–2.3, n = 54) were severely stunted.

### Distribution of double burden of malnutrition by the socio-demographic characteristics

We looked at the distribution of thinness and overweight/obesity by socio-demographic characteristics of the study sample to identify the determinants of nutritional issues in this study sample (Table 3).

**Table 3:** Distribution of thinness and overweight/obesity among adolescents in the Anuradhapura district, Sri Lanka.

| Socio-demographic characteristics | BMI categories |  |  |  |  |  | Statistical Significance |  |
| --- | --- | --- | --- | --- | --- | --- | --- | --- |
|  | Thin |  | Normal |  | Overweight/<br>obese |  |  |  |
|  | n | % | n | % | n | % |  |  |
| Age in years |  |  |  |  |  |  |  |  |
| 13 | 99 | 24.1 | 272 | 66.2 | 40 | 9.7 | Chi-square | 7.487 |
| 14 | 268 | 22.1 | 817 | 67.4 | 128 | 10.6 | p | 0.278 |
| 15 | 252 | 21.2 | 833 | 70.2 | 101 | 8.5 |  |  |
| 16 | 54 | 18.3 | 215 | 72.9 | 26 | 8.8 |  |  |
| **Sex** |  |  |  |  |  |  |  |  |
| Male | 429 | 29.0 | 892 | 60.2 | 160 | 10.8 | Chi-square | 104.92 |
| Female | 244 | 15.0 | 1245 | 76.7 | 135 | 8.3 | p | 0.000<sup>a</sup> |
| **Educational zone** |  |  |  |  |  |  |  |  |
| Anuradhapura | 221 | 20.6 | 733 | 68.3 | 119 | 11.1 | Chi-square | 19.763 |
| Kekirawa | 135 | 26.7 | 321 | 63.4 | 50 | 9.9 | p | 0.011<sup>a</sup> |
| Tambuththegama | 122 | 18.5 | 480 | 72.7 | 58 | 8.8 |  |  |
| Galenbindunuwewa | 89 | 21.9 | 288 | 70.8 | 30 | 7.4 |  |  |
| Kebithigollewa | 106 | 23.1 | 315 | 68.6 | 38 | 8.3 |  |  |
| **School type** |  |  |  |  |  |  |  |  |
| 1AB | 221 | 19.2 | 785 | 68.0 | 148 | 12.8 | Chi-square | 28.846 |
| 1C | 288 | 23.7 | 829 | 68.2 | 98 | 8.1 | p | 0.000<sup>a</sup> |
| Type 2 | 164 | 22.3 | 523 | 71.1 | 49 | 6.7 |  |  |
| **Maternal Education** |  |  |  |  |  |  |  |  |
| Up to grade 11 | 346 | 21.6 | 1122 | 70.0 | 134 | 8.4 | Chi-square | 13.548 |
| O/L and above | 173 | 20.9 | 545 | 66.0 | 108 | 13.1 | p | 0.001<sup>a</sup> |
| **Paternal Education** |  |  |  |  |  |  |  |  |
| Up to grade 11 | 331 | 22.7 | 1009 | 69.2 | 119 | 8.2 | Chi-square | 14.068 |
| O/L and above | 148 | 19.5 | 513 | 67.6 | 98 | 12.9 | p | 0.001<sup>a</sup> |
| **Mother employed** |  |  |  |  |  |  |  |  |
| Yes | 233 | 21.5 | 736 | 67.8 | 116 | 10.7 | Chi-square | 2.51 |
| No | 434 | 21.8 | 1380 | 69.3 | 178 | 8.9 | p | 0.285 |
| **Father employed** |  |  |  |  |  |  |  |  |
| Yes | 602 | 21.4 | 1935 | 68.8 | 277 | 9.8 | Chi-square | 6.013 |
| No | 61 | 27.7 | 144 | 65.5 | 15 | 6.8 | p | 0.049 |
| **Social class** |  |  |  |  |  |  |  |  |
| High | 37 | 18.7 | 129 | 65.2 | 32 | 16.2 | Chi-square | 14.874 |
| Middle | 525 | 21.6 | 1682 | 69.4 | 218 | 9.0 | p | 0.005<sup>a</sup> |
| Low | 73 | 24.6 | 204 | 68.7 | 20 | 6.7 |  |  |

<sup>a</sup> The Chi-square statistic is significant at the 0.05 level.

There was high prevalence of thinness in Kekirawa compared to other areas, while the overweight/ obesity was highest among urban areas (Anuradhapura). A significantly higher percentage of adolescents in type 1AB schools (larger and usually located in urban/semi urban settings) had overweight and obesity (12.8% compared to 8.1% and 6.7% in type 1C and type 2 schools). Students from families with higher parental education level, were significantly more overweight/ obese. The highest prevalence of overweight/ obesity was reported among adolescents from high social class (16.2%).

### Comparison of prevalence of obesity derived from different definitions

To describe the variation of estimates using different definitions, we used WHO growth references, IOTF (international and Asian) and Indian thresholds for BMI in this study sample (Table 4).

**Table 4:** Prevalence of overweight and obesity among 3105 adolescents from rural Sri Lanka by different definitions.

| Age<sup>a</sup><br>(Years) | Prevalence of overweight (95 confidence interval) by the definition used |  |  |  | Prevalence of obesity (95 confidence interval) by the definition used |  |  |  |
| --- | --- | --- | --- | --- | --- | --- | --- | --- |
|  | WHO | IOTF<br>Internati<br>onal | IOTF<br>Asian | Indian | WHO | IOTF<br>Internati<br>onal | IOTF<br>Asian | Indian |
| **Boys** |  |  |  |  |  |  |  |  |
| 13 | 9.1<br>(5.8-14.2) | 8.1<br>(4.6-13.0) | 11.3<br>(7.1-16.7) | 10.8<br>(6.7-16.1) | 3.8<br>(1.8-7.6) | 2.2<br>(0.8-5.4) | 5.4<br>(3.0-9.6) | 6.5<br>(3.7-10.9) |
| 14 | 7.0<br>(5.2 - 9.4) | 6.3<br>(4.4-8.5) | 7.6<br>(5.6-10.1) | 6.8<br>(4.9-9.1) | 3.8<br>(2.5-5.7) | 2.3<br>(1.3-3.8) | 5.7<br>(4.1-8.0) | 7.0<br>(5.2-9.4) |
| 15 | 8.3<br>(6.3-10.8) | 7.1<br>(5.1-9.5) | 8.8<br>(6.6-11.4) | 7.3<br>(5.3-9.7) | 2.4<br>(1.4-4.0) | 1.9<br>(1.0-3.5) | 4.7<br>(3.2-6.8) | 6.2<br>(4.5-8.5) |
| 16 | 6.3<br>(3.3-11.5) | 5.6<br>(2.4-10.7) | 9.8<br>(5.5-15.9) | 9.8<br>(5.5-15.9) | 2.1<br>(0.7-6.0) | 2.1<br>(0.7-6.0) | 2.1<br>(0.7-6.0) | 2.1<br>(0.7-6.0) |
| **Total** | **7.7**<br>**(6.4 - 9.2)** | **6.8**<br>**(5.5-8.2)** | **8.8**<br>**(7.4-10.3)** | **7.8**<br>**(6.5-9.2)** | **3.1**<br>**(2.3-4.1)** | **2.1**<br>**(1.5-3.0)** | **4.9**<br>**(3.9-6.2)** | **6.2**<br>**(5.0-7.5)** |
| **Girls** |  |  |  |  |  |  |  |  |
| 13 | 4.9<br>(2.8 - 8.5) | 4.4<br>(2.2-8.0) | 9.8<br>(6.2-14.4) | 7.6<br>(4.5-11.8) | 2.2<br>(1.0-5.1) | 0.9<br>(0.2-3.2) | 4.0<br>(2.1-7.4) | 3.1<br>(1.5-6.3) |
| 14 | 8.5<br>(6.5-10.9) | 7.5<br>(5.6-9.8) | 12.4<br>(9.9-15.2) | 11.7<br>(9.3-14.5) | 1.9<br>(1.1-3.3) | 1.4<br>(0.7-2.7) | 4.1<br>(2.8-6.0) | 3.4<br>(2.3-5.2) |
| 15 | 5.1<br>(3.6 - 7.1) | 4.9<br>(3.3-7.0) | 10.3<br>(8.0-13.0) | 8.7<br>(6.6-11.2) | 1.3<br>(0.7-2.6) | 1.3<br>(0.7-2.6) | 3.3<br>(2.1-5.0) | 3.1<br>(2.0-4.5) |
| 16 | 7.9<br>(4.6-13.3) | 7.9<br>(4.1-13.4) | 13.8<br>(8.8-20.3) | 11.2<br>(6.7-17.3) | 1.3<br>(0.4-4.7) | 0.7<br>(0.1-3.6) | 2.6<br>(1.0-6.6) | 2.6<br>(1.0-6.6) |
| **Total** | **6.7**<br>**(5.5 - 8.0)** | **6.2**<br>**(5.0-7.4)** | **11.4**<br>**(9.9-13.0)** | **10.0**<br>**(8.6-11.5)** | **1.7**<br>**(1.1-2.4)** | **1.2**<br>**(0.8-1.9)** | **3.6**<br>**(2.8-4.6)** | **3.2**<br>**(2.4-4.2)** |

<sup>a</sup>Age group 13 includes 13–13.9-year-olds, age group 14 includes 14–14.9-year-olds, age group 15 includes 15–15.9-year-olds and age group 16 includes 16–16.9-year-olds

The lowest prevalence for overweight and obesity was seen with IOTF-international thresholds. Especially in girls, IOTF-Asian and Indian thresholds produced higher prevalence of overweight and obesity than WHO and IOTF-international thresholds. McNemar’s test revealed that the prevalence estimates of boys and girls using all 5 definitions were significantly different from each other (p < 0.01) with the exception of difference between Indian threshold and IOTF-Asian threshold in boys (p = 0.5).

### Waist circumference as a marker of obesity

Since this was a large-scale study, WC measurement without clothes was not feasible. Instead, we calculated a correction factor for school uniform in a subsample. Paired sample t-test revealed a significant difference between the waist circumference measurements with and without uniforms in both sexes; mean difference (CI) = 1.04 (0.87 – 1.21), p = 0.000 for boys and 1.61 (1.33 – 1.88), p = 0.000 for girls. Therefore, a correction of 1.04 cm and 1.61 cm was applied to the waist circumference measurements of boys and girls respectively, to get actual waist circumference (without school uniform). BMI had a strong positive correlation with both WC and WHtR in both sexes; r = 0.944 and 0.898 for boys and 0.900 and 0.894 for girls respectively.

To describe the variation of estimates of central obesity using different definitions, we used Indian, South Indian, British WC thresholds and WHtR in this study sample (Table 5).

**Table 5:** Prevalence of central obesity among 3105 adolescents from rural Sri Lanka by different threshold values.

| Age<sup>a</sup><br>(years) | N | The definition used<sup>b</sup> |  |  |  |
| --- | --- | --- | --- | --- | --- |
|  |  | WC-Indian | WC-South Indian | WC-British | Waist to height ratio |
| Boys |  |  |  |  |  |
| 13 | 186 | 5.9 (3.3-0.3) | 11.8 (8.0-17.3) | 12.4 (8.4-17.9) | 8.6 (5.4-13.5) |
| 14 | 574 | 6.3 (4.6-8.6) | 9.2 (7.1-11.9) | 9.0 (6.7-11.3) | 6.8 (5.0-9.2) |
| 15 | 578 | 5.7 (4.1-7.9) | 9.5 (7.4-12.2) | 9.0 (7.0-11.6) | 8.0 (6.0-10.4) |
| 16 | 143 | 4.2 (1.9-8.9) | 8.4 (4.9-14.1) | 6.3 (3.3-11.5) | 6.3 (3.3-11.5) |
| Total | 1481 | 5.8 (4.7-7.1) | 9.6 (8.2-11.2) | 9.2 (7.7-10.6) | 7.4 (6.2-8.9) |
| Girls |  |  |  |  |  |
| 13 | 225 | 2.7 (1.2-5.7) | 6.2 (3.7-10.2) | 8.9 (5.8-13.3) | 4.9 (2.8-8.5) |
| 14 | 639 | 3.0 (1.9-4.6) | 5.9 (4.4-8.1) | 12.4 (10.0-15.1) | 5.6 (4.1-7.7) |
| 15 | 609 | 2.5 (1.5-4.0) | 4.1 (2.8-6.0) | 11.3 (9.0-14.1) | 4.6 (3.2-6.6) |
| 16 | 152 | 2.6 (1.0-6.6) | 2.6 (1.0-6.6) | 11.2 (7.1-17.2) | 7.9 (4.6-13.3) |
| Total | 1625 | 2.6 (2.0-3.6) | 5.0 (4.0-6.2) | 11.4 (9.9-13.0) | 5.3 (4.4-6.6) |

<sup>a</sup> Age group 13 includes 13–13.9 year olds, 14 includes 14–14.9 year olds, 15 includes 15–15.9 year olds and 16 includes 16–16.9 year olds
<sup>b</sup> Suggested thresholds: Indian- 70<sup>th</sup> percentile, South Indian- 75<sup>th</sup> percentile, British- 90<sup>th</sup> percentile, waist to height ratio- >0.5

Prevalence estimates of central obesity were significantly different between each of the four definitions (McNemar’s test; p < 0.001).

## Discussion

This study is the largest study reported in the literature on double burden of malnutrition among rural adolescents in Sri Lanka. In this study, we showed that the double burden of malnutrition among adolescent school children in rural Sri Lanka is a priority public health issue with 9.5% prevalence of overweight/obesity and 21.7% prevalence of thinness. In addition, 12.8% of children were stunted, showing that the nutrition problem is chronic. We further showed the need for consensus on prevalence estimates, which could widely vary when using different thresholds.

Within country comparison of obesity data alone, our prevalence estimates seemed lower. In Colombo, the capital of Sri Lanka, prevalence of obesity was reported as 5.8% among boys and 8.6% among girls using the similar WHO thresholds [25]. Even in a 2004 study done in Colombo among 8-12 years old, 4.3% boys and 3.1% girls were reported to be obese. However, the latter study was using IOTF-international thresholds [11]. Our comparable figures using the same thresholds were still lower (Table 4). In a recent study among preadolescents (5-11 years) in the North Central province, where the Anuradhapura district is located, the prevalence of obesity using similar WHO thresholds was reported as 2.5% [26]. Although, the prevalence of overweight was lower in our study compared to a 2004 study in urban Colombo [11], it was higher than prevalence reported from other parts of the country, including an all island study. In the urban Colombo study, prevalence of overweight was 11.1% in boys and 10.8% in girls using the IOTF thresholds (international) [11]. Using WHO thresholds, a 1997 study in Colombo (10) gave a prevalence of overweight as 4.0% and a 2008 study in Kaluthara (a district in urban western province) gave prevalence of overweight as 4.4% among boys and 8.7% among girls [9]. Prevalence of overweight was 1.7% among boys and 2.7% in girls in an all island study in 2006 when using IOTF international threshold [27]. The recent study carried out in Anuradhapura district gave quite lower prevalence of overweight (3.0%). However, the study was done among under-privilege schools excluding many semi-urban schools in the district that may have given this low prevalence. They also have selected preadolescent children and have shown that prevalence of overweight increased with age [26].

Reported prevalence of obesity lies around 1-2% in South Asian countries including India, while prevalence of thinness is around 22.7-30.7% in India [2]. Hence, our comparative data (with international thresholds) for obesity and thinness lie within the reported South Asian prevalence estimates. Increased prevalence of overweight in this rural district may be a harbinger for complex social, cultural and economic factors that drive change in food habits and physical activity. Most importantly, well documented social disparities are clearly playing a significant role in malnutrition in this rural community. Children of educated, high social class families were having a higher prevalence of overweight and obesity. Low social class also clearly showed higher rates of prevalence of under nutrition. A similar pattern has been reported in previous studies in Sri Lanka where higher prevalence of overweight and obesity has increased with family income and thinness being high among low socioeconomic group [28,29]. Rising prevalence of overweight in this rural community may herald an epidemic of obesity. There is no reason to say that this is not the situation in many rural settings in Sri Lanka, except few places, where the under-nutrition is still the priority. The behavioral as well as policy interventions to control overweight and obesity at population level usually take years to see an impact. It is of utmost importance to include policy interventions which could guide behavioral changes to halt the increasing trend of overweight and obesity in these communities.

While the new threats due to overweight and obesity are emerging, the known issues of under-nutrition are yet to be addressed in rural Sri Lanka. The reason for boys to become more thin, fat and short than girls needs further exploration. Higher prevalence of malnourishment among boys is a finding which agrees with many studies done in Sri Lankan as well as other Asian populations [9,11,30,31]. Increasing prevalence of overweight and obesity coexisting with under nutrition (thinness and stunting) had been reported in Asian countries, in low income groups and nutrition deficit regions [32]. The underlying behavioral or socio-cultural practices for this issue as well as high and gradually increasing prevalence of under-nutrition from under five children to adolescents need to be investigated with a fresh approach, because the strategies we have been using over last few decades seems not working.

In this study, we clearly showed that scientific approaches are needed for developing country specific threshold values to determine obesity, especially as determinants of future adverse health outcomes.

## Conclusions

High incidence of overweight in a rural district shown by our study warrants revision of school health program to address the emerging issue of obesity as well as to include new approaches to combat age old challenge of malnutrition. Prospective studies as well as consensus is required to as what standards are to be used in Sri Lanka for clinical practice and public health interventions. This may be true for many countries in South Asia as well as other parts in the world, where the genetic risks are different.

## Data Availability

Data file is uploaded with the manuscript as supportive information Also available at https://docs.google.com/spreadsheets/d/e/2PACX-1vRfCxDhLOCqmNUgcxJ6osRP78nVMMLEfFeTe5D8tJg3vS-8Xo4dOvBsONTLf8nlKQ/pubhtml

## Acknowledgements

We would like to thank K. Bopeththa, J. Chandima, temporary assistant lecturers affiliated to faculty of Medicine and Allied Sciences, Rajarata University of Sri Lanka, SMAGB Senevirathna and GLL Indrani for their participation in data collection and data entering processes.

## Notes

### Competing Interest Statement

Authors BLG, NSK and SS declare no conflicts of interest. Author SBA is a member of the editorial board of the Journal PLOS Global Public Health.

### Funding Statement

Rajarata University of Sri Lanka, Mihintale, Sri Lanka (http://www.rjt.ac.lk/) awarded two research grants (RJT/RP&HDC/2012/Med. & Alli. Sci. /R/05 and RJT/RP&HDC/2013/Med. & Alli. Sci. /R/04) to BLG. The funders had no role in study design, data collection and analysis, decision to publish, or preparation of the manuscript.

### Author Declarations

Ethics review committee of the faculty of Medicine and Allied Sciences, Rajarata University of Sri Lanka, Mihintale, Sri Lanka.

